# Spatial simulation of COVID-19 new cases development

**DOI:** 10.1101/2021.09.19.21263799

**Authors:** Isaac Chen, F. Liu

## Abstract

The time dependent SIR model is extended to simulate infection across spatial boundaries. We used New Jersey data as an example to test the extended SIR model. Infection from neighboring counties are modelled by connectivity matrix where each pair of neighboring counties has an element in the connectivity matrix. The magnitude of this matrix element represents the degree to which the infected from one county can affect the susceptible in one of its neighboring counties. Simulated result from the extended spatial SIR model is compared with observed new COVID-19 cases measured in the 21 counties in New Jersey. The extended model has to solve 84 simulated functions simultaneously and the large number of parameters involved in the spatial SIR model are auto tuned using genetic algorithm.

## 1 Introduction

SIR model [1] [2] has been used extensively in modelling COVID-19 [3] [4] [5] [6] [7], [8], [9]. Most of the work focus on comparing the measured and simulated new cases and evaluating the effect of vaccination and quarantine effect. The primitive SIR model has three groups of population that are coupled with each other: susceptible, infected and recovered. Work done by [5] has expanded the SIR model to include mortality effect due to the more dangerous nature of COVID-19.

An aspect of COVID-19 modelling that has not been extensively studied is how it can spatially spread from countries to countries, states to states, and cities to cities. This is due to the formulation of the primitive and extended SIR model being differential equations with time dependence only [1] [10]. Studies investigating spatial transmission focus on observations [11], [12], [13]. Few work [14] focuses on modelling spatial transmissions of COVID-19.

To model the spatial spread of COVID-19 across area boundaries, we extend the ordinary differential equations to partial differential equations to include a spatial diffusion term due to uneven proportion of infection across spatial boundaries. The SIR populations of neighboring spatial regions are coupled through the diffusion term. In such an approach, the SIR populations are functions of both time and space. These functions are solved from the set of partial differential equations.

In this study, we present how a spatially connected SIR model is used to model the COVID-19 cases observed in the 21 counties in New Jersey, US. We also discuss how genetic algorithm is used to guide the auto tuning process for the large number of parameters involved in the equations to find the solution of the spatially connected SIR model equations.

## 2 SIR Model with spatial connection

We start with the extended SIR model [5] for a single spatial location.

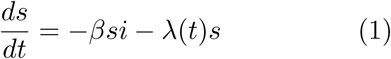

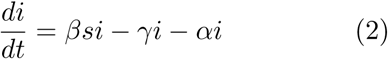

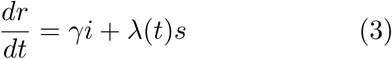

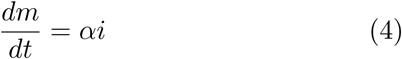

where *s, i, r, m* represents total population scaled susceptible, infected, recovered, and mortality proportions; *β* is the coupling coefficient between local susceptible and infected populations dependent on the infection rate of COVID-19; *λ*(*t*) is a time dependent control measure function that transfers susceptible to recovered population; *γ* represents the rate at which infected population can recover; finally *α* represents the mortality rate of the infection.

With the introduction of spatial dependence, the SIRM populations become both time and spatial dependent, therefore the equations become partial differential equations.

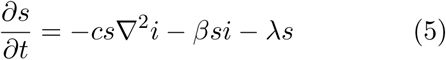

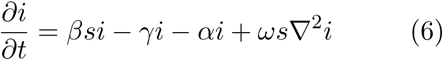

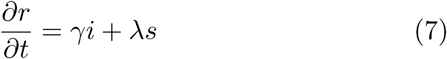

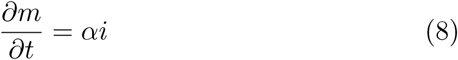

where *c* represents diffusion rate in between connected spatial regions and *s*∇^2^*i* is the coupled term representing rate of change in infection due to difference in infection proportion in population between the spatial location and its neighboring locations.

Because of the discrete nature of spatial locations that have been artificially designated, e.g. countries, states, and counties. The spatial connection term *cs*∇^2^*i* can be represented using matrix approach. Each matrix element describes connectivity between spatial regions that can affect each other.

To make things more concrete, we applied the extended model in New Jersey that has 21 counties. The connectivity matrix therefore has 21 by 21 elements. But not all elements has non-zero values. In this work, only counties neighboring each other has a non-zero element in the matrix representation. Ignoring exchange of infected population from outside of New Jersey, we use 84 functions to represent the *s, i, r, m* variables for 21 counties in New Jersey. We discretize the equations into the following form,

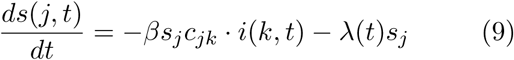

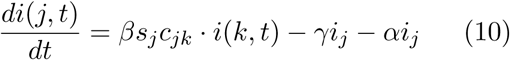

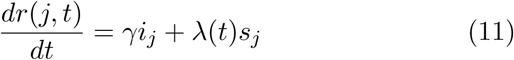

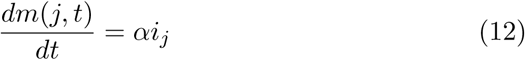

*j* is the index of the county that runs from 1 to 21. *c*_*jk*_ is the spatial connection matrix that represents the propagation coefficient across county boundaries between the current (*j*-th) county and *k*-th county.

The term *c*_*jk*_ · *i*(*k, t*) should be understood as a vector multiplication,

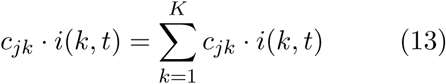

where *K* is the total number of counties, in this case, 21 for New Jersey. Note that the diagonal elements of the connection matrix *c*_*jj*_ ≡ 1 when *k* = *j*. Because each diagonal element of the connection matrix represents the coupling between the *s* and *i* function of the county itself. The summation represents the effective number of infected cases that can infect susceptible population in the *j*-th county.

Note that the control measure term *λ*(*t*) is a time dependent function [5] formulated after the sigmoid function,

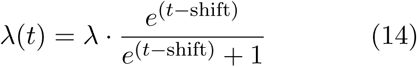

The step wise shape of the sigmoid function allows the model to turn on control measures such as quarantine, vaccination etc at time specified by the ‘shift’ parameter.

## 3 Auto tuning SIR parameters

Due to the large number of parameters involved in the 84 differential equations including the elements of the connection matrix, it is not feasible to manually adjust all the parameters involved. We adopt the same approach in [6] to use genetic algorithm to auto tune the following parameters, *α, β, γ, λ*, shift, *c*_*j,k*_ for each county in New Jersey.

First we define the residual function as

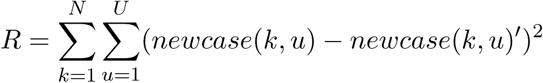

where *k* represents the county, *u* represent the number of days from pandemic outbreak, new-case(k,u) is from observation, and newcase(k,u)’ is from model simulation.

Our goal is to minimize the function such that simulation can match the observation as close as possible. To minimize the residual, we will fine tune all the parameters used in SIR model through genetic algorithm.

The models is set up to start with an initial set of parameters for all the counties shown in Table 1.

**Table 1:** Default parameter for the spatial SIRM solver

| Parameter | Default Value |
| --- | --- |
| $\alpha$ | 1e-6 |
| $\beta$ | 1.5 |
| $\gamma$ | 0.1/14 |
| $\lambda$ | 0.05 |
| $c_{j,k}$ | 0.05 |
| shift | 90 (days) |

During each iteration, the parameters are mutated (updated) randomly (Equation 15) to form a new genetic code. The mutated genetic code is used to configure the 84 differential equations for New Jersey, simulating new cases in each county by solving the 84 equations simultaneously.

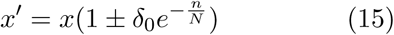

where *x*′ is the new genetic code which could be any of the parameters used in the simulation; *x* is the current genetic code; *δ*_0_ is the seed of mutation; 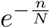 is the decay term that reduces the magnitude of mutation over time

After each iteration of simulation for 180 days after the onset of COVID-19 in New Jersey, the simulated new cases is used in the residual function. If the residual decreases (Figure 1), the mutated genetic code replaces the previous genetic code. Otherwise, the mutated genetic code is discarded and the previous code is used to start a new iteration of simulation.

**Figure 1:**
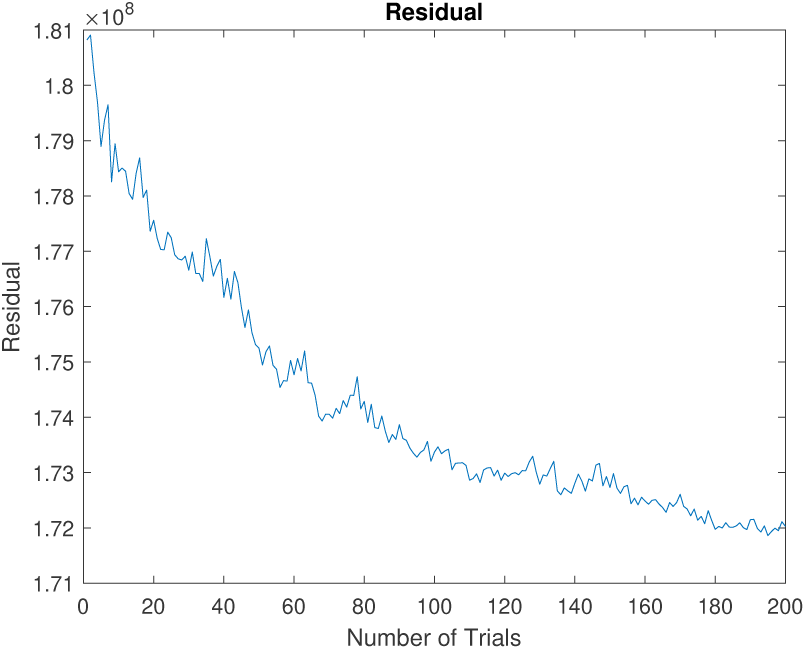
The residual of the SIR Model getting lower as the genetic algorithm optimizes the variables to better fit the collected data.

This process allows the algorithm to harness the computing power of the computers and slowly but surely the residual decreases over generations of genetic mutation as shown in Figure 1. The decrease in residual means better agreement between simulated model result compared with observations.

## 4 Result

Using the SIR model, it is possible to use differential equations and a genetic algorithm to simulate the propagation of a virus. The generated curves, after going through many rounds of optimizations by the genetic algorithm, are able to closely match the data provided by Johns Hopkins for the number of cases in each county every day.

The observed new cases tend to fluctuate significantly (Figure 4) which creates difficulty for the residual calculation. Because the residual calculation uses the difference between observed daily new cases and predicted daily new cases, the fluctuation in the observed new cases can deviate from the smooth predicted curve significantly and artificially increase the residual. Therefore a rolling average smoothing is applied to the observed new cases data points to reduce such artificial cause of residual difference. Another benefit of smoothing the observed daily new cases helps to remove outliers, such as a day with 0 recorded cases among days with hundreds of them. Removing such outliers also reduces calculated residual.

The genetic algorithm is able to create a simulated virus propagation curve for new cases each day and optimize it to match the number of new cases for each county as closely as possible. The degree to which they match is measured by the residual. The smaller the residual the better the agreement between the observation and the prediction. Residual between observation and prediction for the first 200 trials in Figure 1.

To reach good agreement between observation and prediction, large number of iterations (generations of mutation) is needed. Figure 2 shows how the observed (blue) matches the numerical prediction (red) after 4000 iterations.

**Figure 2:**
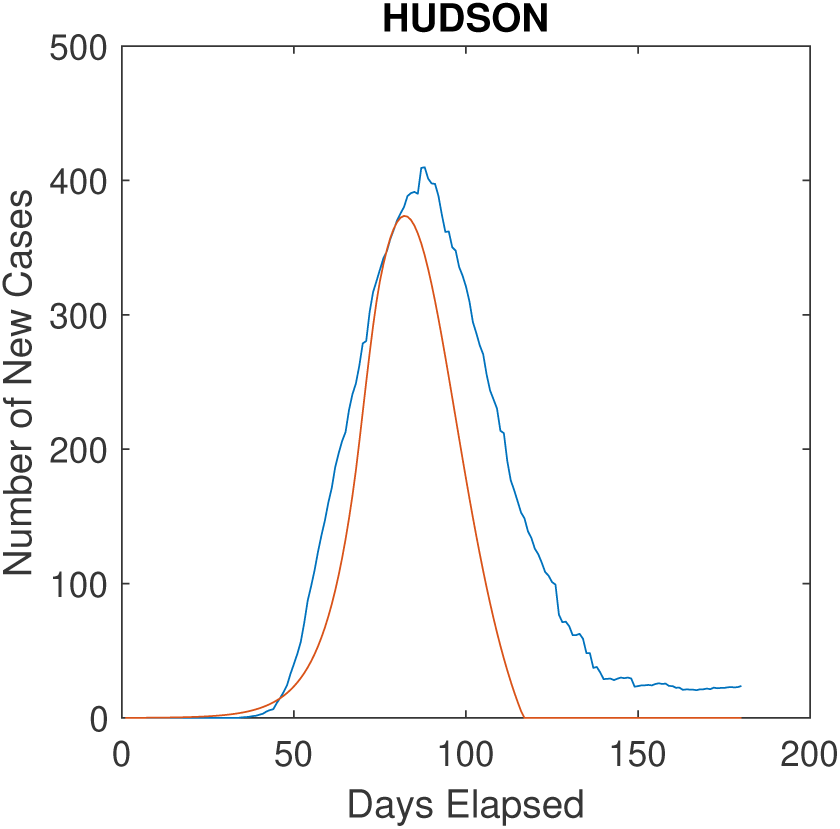
The SIR Model created curve (red) overlaid on top of recorded new cases (blue) for Hudson County, NJ

Counties with large observed new cases tend to show good agreement between observation and prediction such as Hudson (Figure 2) and Passaic (Figure 3). Table 2 and Table 3 show the parameters generated from genetic algorithm used to produce the close agreement for those two counties. These numbers can be compared with Table 1 to examine how the parameters have evolved by minimization of the residual through genetic algorithm.

**Table 2:** Values of genetic variables for Hudson County after solving

| Parameter | Value |
| --- | --- |
| $\alpha$ | 8.2932e-07 |
| $\beta$ | 1.3910 |
| $\gamma$ | 0.0068 |
| $\lambda$ | 0.0417 |
| Bergen Affecting Hudson | 0.0443 |
| Passaic Affecting Hudson | 0.0541 |
| Essex Affecting Hudson | 0.0719 |
| shift | 71 (days) |

**Table 3:** Values of genetic variables for Passaic County after solving

| Parameter | Value |
| --- | --- |
| $\alpha$ | 8.2967e-07 |
| $\beta$ | 1.7601 |
| $\gamma$ | 0.0063 |
| $\lambda$ | 0.0462 |
| Bergen Affecting Passaic | 0.0379 |
| Essex Affecting Passaic | 0.0410 |
| Morris Affecting Passaic | 0.0407 |
| Sussex Affecting Passaic | 0.0318 |
| shift | 85 (days) |

**Figure 3:**
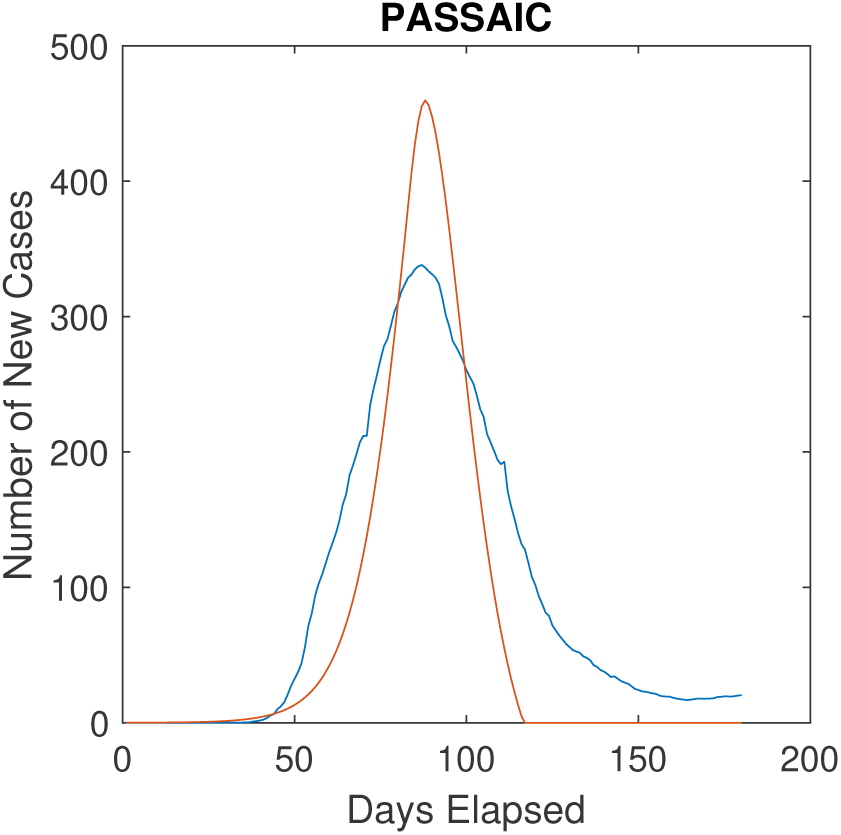
Graph (red) overlaid on collected data (blue) for Passaic County, NJ (Specify the final genetic information used for the result.)

## 5 Discussion

The new case patterns in the 21 counties of New Jersey since the onset of the COVID-19 pandemic are modelled by solving discrete differential equations coupled through a connectivity matrix to simulate spatial connection. The elements of the connectivity matrix represents the influence of infection from the neighbouring counties. Due to the large number of parameters involved, genetic algorithm is used to fine tune the relevant parameters used in the modelling.

One noticeable artifact of the genetic algorithm is that it prioritizes optimizations for counties with higher numbers of cases, since this optimization will lower the residual more dramatically. However, this neglects optimizations on counties with fewer cases (See Figure 4). To alleviate this, we separated each county into one of three tiers, which each tier corresponding to a range of cases. By allowing the genetic algorithm to optimize counties within each tier separately, it is able to first focus on counties with high cases and then work separately on counties with fewer cases.

**Figure 4:**
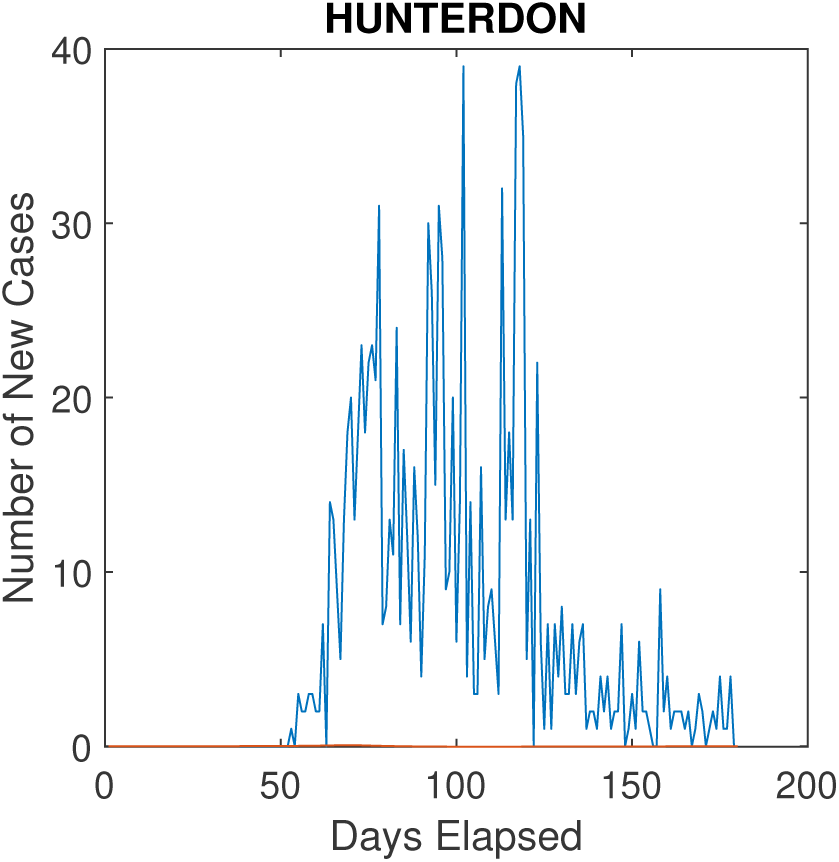
The genetic algorithm (red) has not optimized the residual for Hunterdon County well because it has very few cases (at max 40 per day). Also note the importance of smoothing the data to remove outliers with 0 cases.

Another problem with the genetic algorithm is its inability to lower the residual past a certain point. Once it reaches this point, further generations will create residuals that hover around approximately the same value. For our study with the 21 counties in NJ, this happens around 2 ∗ 10^7^. This suggests a local stationary point in the space spanned by the parameters used by the algorithm dominated by the high new cases numbers from certain counties such as Passaic (Figure 3).

The genetic algorithm is sometimes unable to change the variables enough to lower the residual drastically, requiring human intervention. We may have to manually change certain variables for certain counties to get their graphs to match with provided data. However, ideally we would never have to intervene and the algorithm would process everything automatically, because human intervention can disrupt the results of other counties due to the spatial connection aspect we implemented. This makes the experiment less applicable to the natural situation of the propagation of a virus.

The spatial connection aspect that was implemented works quite well. Counties that are directly adjacent to others with large proportions of cases tend to also have larger proportions of cases, simulating infected people who traveled between the counties. However, it is still not perfect, as certain counties seemed to be unaffected by neighboring counties, even with drastic changes in the spatial coefficient that affects the number of people traveling between them. This is most pronounced in counties with lower population, as the genetic algorithm tends to favor counties with higher population even within each population tier in our tiered model.

With more optimizations in code and a better understanding of initial variables, better results may be produced. This method allows for great flexibility, such as changes in government regulations, because they can be reflected by simply changing a variable. Now, it is important to quantify the effect of changes in human behavior versus the change in the variables. With better insight into human behavior in the virus, it becomes possible to roughly estimate the impact of a virus in the future, and plans can be made accordingly.

## Data Availability

Data can be made available upon request.

https://github.com/CSSEGISandData/COVID-19

## References

[1] Kermack, W.O.; McKendrick, A.G. (1927). “A Contribution to the Mathematical Theory of Epidemics”. Proceedings of the Royal Society A. 115 (772): 700–721. Bibcode:1927RSPSA.115..700K. doi:10.1098/rspa.1927.0118.

[2] David Smith and Lang Moore, “The SIR Model for Spread of Disease”, Convergence (December 2004)

[3] Matteo Chinazzi, et al, “The effect of travel restrictions on the spread of the 2019 novel coronavirus (COVID-19) outbreak”, Science, 24 Apr 2020, Vol 368, Issue 6489, pp. 395–400, DOI: 10.1126/science.aba9757

[4] Li Qun, et al, “Early Transmission Dynamics in Wuhan, China, of Novel Coronavirus–Infected Pneumonia”, N Engl J Med 2020; 382:1199–1207, DOI: 10.1056/NEJ-Moa2001316

[5] Lee, C. and Liu, F. “Effect of control measure on the development of new COVID-19 cases through SIR model simulation”, medRxiv 2020.10.27.20220590

[6] Lee, K. and Liu, F. “Auto tuning SIR model parameters using genetic algorithm”, medRxiv 2021.07.19.21260722

[7] Copper I., et al. (2020), “A SIR model assumption for the spread of COVID-19 in different communities”, Chaos, Solitons and Fractals 139 (2020) 110057

[8] Giordano, G.; Blanchini, F.; Bruno, R.; Colaneri, P.; Di Filippo, A.; Di Matteo, A.; Colaneri, M. (2020). “Modelling the COVID-19 epidemic and implementation of population-wide interventions in Italy”. Nat Med 26, 855–860 (2020).

[9] Riou J, Althaus CL. “Pattern of early human-to-human transmission of Wuhan 2019 novel coronavirus (2019-nCoV), December 2019 to January 2020. Euro Surveill. January 20, 2020 https://www.ncbi.nlm.nih.gov/pmc/articles/PMC7001239/

[10] Ashlynn R. Daughton, NicholasGenerous, Reid Priedhorsky & Alina Deshpande received: 03 October 2016 accepted: 28 February 2017 Published: 18 April 2017

[11] Zheng R, Xu Y, Wang W, Ning G, Bi Y. Spatial transmission of COVID-19 via public and private transportation in China. Travel Med Infect Dis. 2020;34:101626. doi:10.1016/j.tmaid.2020.101626

[12] Liu, J., Zhou, Y., Ye, C. et al. The spatial transmission of SARS-CoV-2 in China under the prevention and control measures at the early outbreak. Arch Public Health 79, 8 (2021). https://doi.org/10.1186/s13690021-00529-z

[13] Rader, B., Scarpino, S.V., Nande, A. et al. Crowding and the shape of COVID-19 epidemics. Nat Med 26, 1829–1834 (2020). https://doi.org/10.1038/s41591-020-1104-0

[14] Danon, L. et al., 2021 A spatial model of COVID-19 transmission in England and Wales: early spread, peak timing and the impact of seasonalityPhil. Trans. R. Soc. B3762020027220200272 http://doi.org/10.1098/rstb.2020.0272

